# Systematic review and meta-analysis of intraventricular antibiotics for neonatal meningitis and ventriculitis

**DOI:** 10.1101/2023.09.07.23295218

**Authors:** Doriam Alejandrino Perera Valdivia, Edgar Abraham Herrera Pérez, Luis Roberto Zapata Vega, José Miguel Hurtado García, Karen Vanessa Herrera

## Abstract

**Background:** Many pediatric and neurosurgical studies have been published regarding intraventricular antibiotics in neonatal meningitis and ventriculitis. We aimed to determine the safety and effectiveness of intraventricular antibiotics in neonates with meningitis and/or ventriculitis and analyze the quality of available evidence.

**Methods:** We systematically reviewed scientific literature from the PubMed, EMBASE, LILACS, and SCOPUS databases. Randomized experimental and observational studies were included. The Cochrane methodology was used for systematic reviews.

**Results:** Twenty six observational studies and one randomized clinical trial involving 272 patients were included. The risk of bias in both pediatric and neurosurgical studies was high, and the quality of evidence was low (evidence level C). In the pediatric studies, no significant differences in mortality were found between intraventricular antibiotics and only systemic antibiotic [25.4% vs 16.1%, OR=0.96 (0.42– 2.24), *P*=0.93]. However, when analyzing the minimum administered doses, we found a lower mortality when a minimum duration of 3 days for intraventricular antibiotics was used compared to only systemic antibiotic [4.3% vs 17%, OR=0.22 (0.07–0.72), *P*=0.01]. In the neurosurgical studies, the use of intraventricular antibiotics in ventriculitis generally results in a mortality of 5% and a morbidity of 25%, which is lower than that in cases where intraventricular antibiotics were not used, with an average mortality of 37.3% and a morbidity of 50%.

**Conclusion:** Considering the low quality of evidence in pediatric and neurosurgical studies, we can conclude with a low level of certainty that intraventricular antibiotics may not significantly impact mortality in neonatal meningitis and ventriculitis. However, reduced mortality was observed in cases treated with a minimum duration of 3 days of intraventricular antibiotic, particularly the multidrug-resistant or treatment-refractory infections. Higher-quality studies are needed to improve the quality of evidence and certainty regarding the use of intraventricular antibiotics for treating neonatal meningitis and ventriculitis.

## INTRODUCTION

Neonatal meningitis (along with its severe forms such as ventriculitis) is a disease with one of the highest mortality and morbidity rates. It has a mortality rate of 40–58% in developing countries and 5–25% in developed countries.^1–5^ Those who survive this condition present with significant morbidity in up to 50% of cases, regardless of geographic location.^6–8^ Worldwide, the mortality from neonatal meningitis and sepsis is estimated to be between 248,000 and 402,000 children per year.^9^

In the 1970s and 1980s, intraventricular antibiotics were successfully used in some cases of neonatal meningitis.^10–14^ In 1980, a randomized clinical trial of intraventricular gentamicin in neonatal meningitis ^15^ reported a very high mortality associated with the use of intraventricular antibiotics, which drastically reduced interest in the clinical use and research of intraventricular antibiotics in neonates, in contrast to the increased clinical use and research of intraventricular antibiotics in adults.

However, in recent years, many observational studies have reported the safe and effective use of intraventricular antibiotics in neonates in both pediatrics and neurosurgery. Therefore, this study aimed to systematically investigate the effectiveness of intraventricular antibiotics in reducing mortality and morbidity in neonatal meningitis and ventriculitis.

Owing to the differences between the studies, we divided the review as follows: a) intraventricular antibiotics in neurosurgery and b) intraventricular antibiotics in pediatric patients. Neurosurgery research included studies on ventriculitis associated with structural disorders (hydrocephalus, myelomeningocele, empyema, etc.) typically related to ventricular medical devices. Research in pediatrics included studies in the pediatric field, primarily focused on treating patients with meningitis and ventriculitis, which were unrelated to structural alterations.

## METHODS

The recommendations of the PRISMA 2020 guidelines and the Cochrane Manual of Systematic Reviews for interventions version 6.3 were followed.^16–17^ Only experimental and observational studies in which management was standard treatment for meningitis or ventriculitis were included.^18^ Regarding the management of ventriculitis, only studies that removed the infected ventricular system and repositioned a new ventricular system upon resolution of ventriculitis after antibiotic therapy were included. Neurosurgery studies included those on the treatment of neonatal meningitis or ventriculitis associated with the use of intraventricular devices (ventriculoperitoneal shunts or external ventricular drains) or associated with neurosurgical resolution of diseases such as intraventricular hemorrhage, hydrocephalus, and myelomeningocele. The electronic search included the following keywords: “ventriculitis,” “intraventricular antibiotic,” “meningitis,” “neonates,” “newborn,” “intrathecal,” “amikacin,” “vancomycin” and “colistin.”

We reviewed the PubMed, EMBASE, LILACS, and SCOPUS, included studies published in any language. The inclusion criteria for the meta-analysis studies included: 1. comparative studies (randomized clinical trials or cohort studies) in which the treatment groups were clearly differentiated (systemic antibiotics only vs. intraventricular antibiotics with systemic antibiotics); 2. the results of the study variables were reported; 3. adequate and current management of meningitis and ventriculitis were used; 4. not greater than 20% of patients were lost to follow-up of the variables of interest; 5. more than 10 participants.

The primary response variable was the mortality rate, while the secondary response variables included infection cure, morbidity, complications, neurological sequelae, and adverse reactions.

The methodological quality and risk of bias for observational and experimental studies were reviewed using the ROBINS-I and ROBINS-II tools, respectively. The search, review, and analysis of the risk of bias of each study were carried out by two authors (DP and KH). In case of any differences in the analyses, a consensus was attempted; if the consensus was not reached, the opinion of a third reviewer (LZ) was requested.

The Review manager 5.4 program was used for the meta-analysis. Relative risks were used for dichotomous variables, each with its confidence interval. We visually reviewed the heterogeneity of the studies and used the Chi-square test for heterogeneity. If significant heterogeneity was found, the possible methodological and clinical causes that could explain it were reviewed and subgroup analysis was performed.

All meta-analysis models were reviewed to obtain the effects of the intervention (fixed effects model, random effects model, and Peto) and to estimate the global effect we used the most conservative model, which is the one that achieved the value closest to zero or had a P-value closest to 1 (least significant). We intended to see the risk of publication bias using the funnel plot if the number of studies for meta-analysis was equal to or greater than 10. These number of studies were not achieved in the meta-analysis; thus, this graph was not produced. A forest plot was used for graphical representation of the meta-analysis.

The GRADE approach recommended by the Cochrane Manual was used to evaluate the certainty of evidence for the most important variables in this review.^17^

In the case of neurosurgery research, as no adequate analytical (comparative) studies were found, the results of the individual studies were combined to obtain a descriptive synthesis.

## RESULTS

Our search yielded a total of 617 studies. After reviewing the titles, abstracts, and some full-text articles, we selected 23 studies for the descriptive analysis and 4 studies for the meta-analysis that include a total of 272 patients (Figure 1).

**Figure 1.**
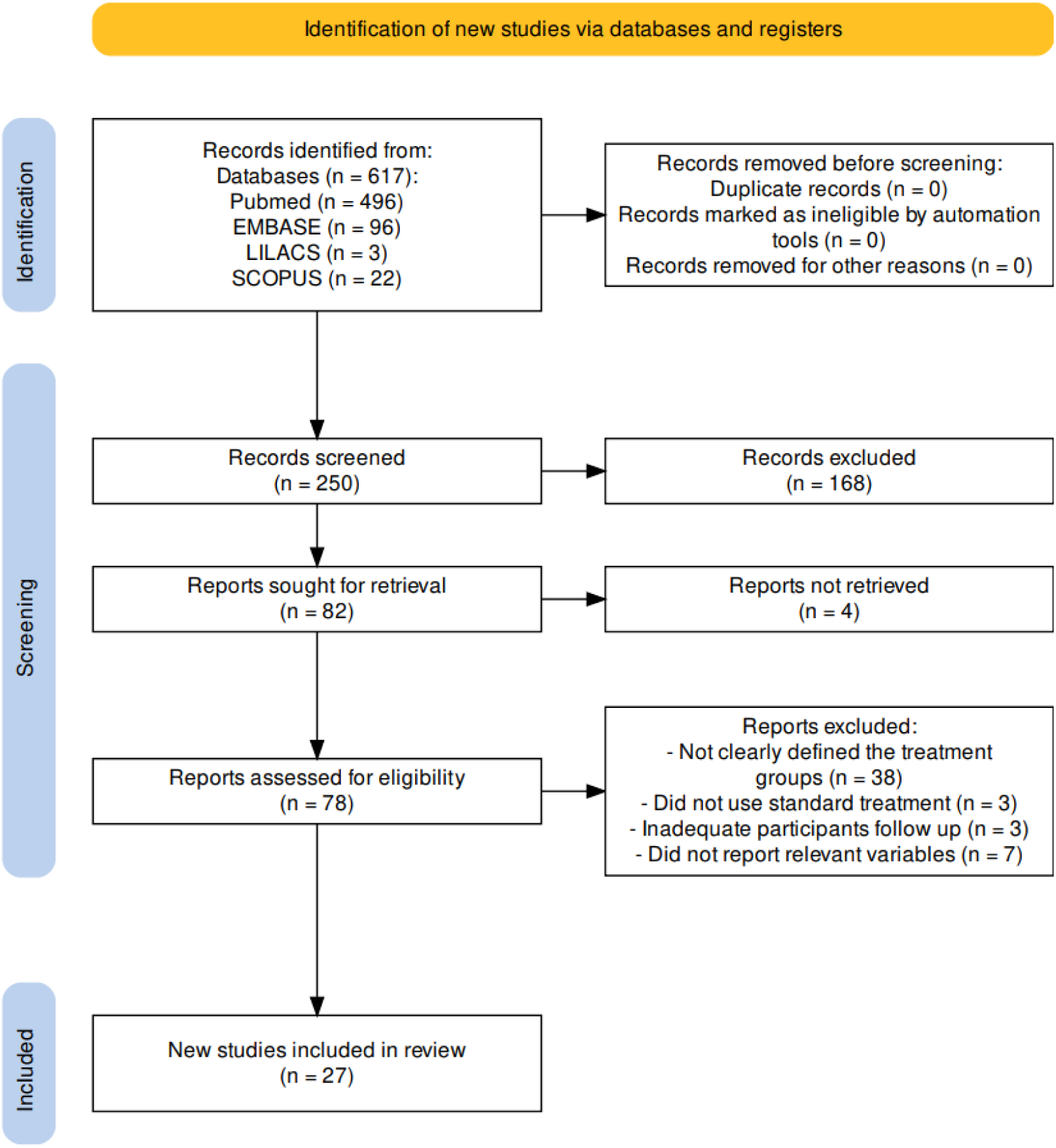
Flowchart of this systematic review. Flowchart generated using PRISMA online software. (37)

The certainty of this estimate was low (downgraded owing to the high risk of bias, imprecision, and publication bias; evidence was upgraded to one level for consistency of results, as most studies reported similar results).

### A) Intraventricular antibiotics in neurosurgery

Our analysis included 19 studies using intraventricular antibiotics in neonates undergoing neurosurgery, comprising 59 patients (Supplement 1). Most of these studies are case reports^19–31^ or small case series^32–35^, with very few small comparative studies (cohorts).^36,10^ Another 26 studies on neonates also used intraventricular antibiotics to treat neuroinfections in neurosurgery, but they were not included in our analysis because the outcomes of interest were not specified in sufficient detail.

A meta-analysis could not be performed as none of the analytical studies (comparative studies) of neurosurgery research reached the appropriate size that we had anticipated in the inclusion criteria.

Table 1 shows the obtained grouped results. Mortality with the use of intraventricular antibiotics in neurosurgery is 5%, and an infection cure is achieved in 83% of cases. In general, the studies hav reported limited information on the morbidity or adverse effects associated with intraventricular antibiotics. The main reasons for their use were refractory central nervous system and multidrug-resistant infections. The main difference between these was that in multidrug-resistant infections, intraventricular antibiotics were used close to the start of treatment, and in refractory infections, intraventricular antibiotics were used when systemic antibiotic therapy had already failed.

**Table 1.**
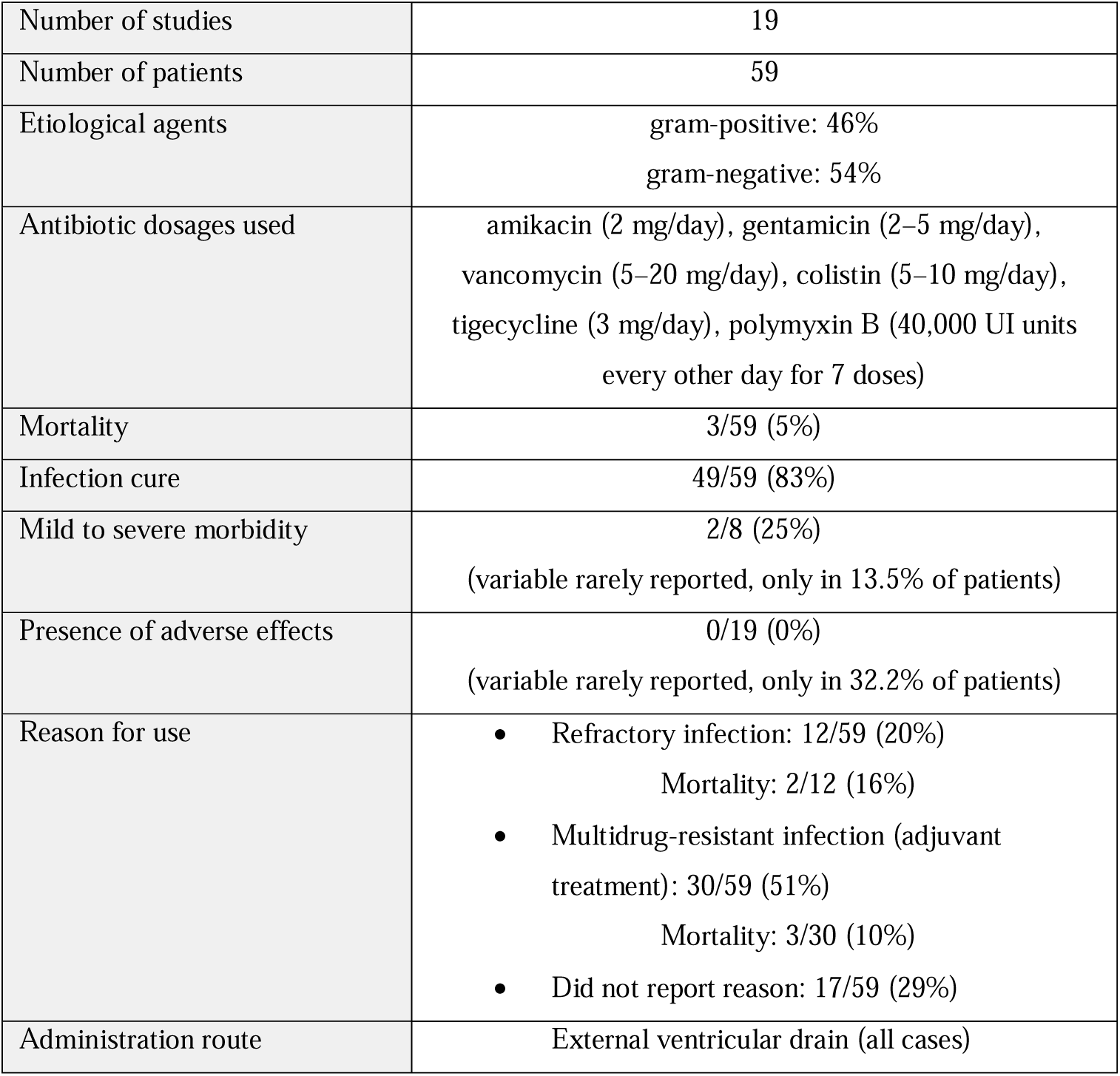
Description of neurosurgical studies using intraventricular antibiotics in neonates.

|  |  |
| --- | --- |
| Number of studies | 19 |
| Number of patients | 59 |
| Etiological agents | gram-positive: 46%<br>gram-negative: 54% |
| Antibiotic dosages used | amikacin (2 mg/day), gentamicin (2–5 mg/day),<br>vancomycin (5–20 mg/day), colistin (5–10 mg/day),<br>tigecycline (3 mg/day), polymyxin B (40,000 UI units<br>every other day for 7 doses) |
| Mortality | 3/59 (5%) |
| Infection cure | 49/59 (83%) |
| Mild to severe morbidity | 2/8 (25%)<br>(variable rarely reported, only in 13.5% of patients) |
| Presence of adverse effects | 0/19 (0%)<br>(variable rarely reported, only in 32.2% of patients) |
| Reason for use | Refractory infection: 12/59 (20%)<br>Mortality: 2/12 (16%)<br>Multidrug-resistant infection (adjuvant treatment): 30/59 (51%)<br>Mortality: 3/30 (10%)<br>Did not report reason: 17/59 (29%) |
| Administration route | External ventricular drain (all cases) |

### B) Intraventricular antibiotics in pediatrics

We included four comparative research studies in a pediatric setting that met our inclusion criteria to perform a meta-analysis.^15,38–41^ These four studies comprise a total of 115 patients.

For the descriptive synthesis analysis, we included 4 observational studies (Supplement 2).^10–12,42^ Table 2 presents a synthesis of these descriptive studies in pediatric research. Table 3 presents the characteristics of the studies included in this meta-analysis. The only two outcome variables amenable to meta-analyzed were mortality and neurological morbidity (Table 4).

**Table 2.** Description of observational pediatric studies using intraventricular antibiotics in neonates. *dose not specified.

|  |  |
| --- | --- |
| Number of studies | 4 |
| Number of patients | 26 |
| Etiological agents | gram-positive: 3.8%<br>gram-negative: 96.1% |
| Antibiotic dosages used | colistin*, rifampicin (5 mg/day), gentamicin (1- 2 mg/day) ,<br>chloramphenicol* |
| Mortality | 2/26 (7.7%) |
| Infection cure | 24/26 (92.3%) |
| Mild to severe morbidity | 5/18 (27.7%) |
| Presence of adverse effects | 5/16 (31.2%) |
| Reason for use | Refractory infection: 15/26 (57.7%)<br>Mortality: 2/15 (13%)<br>As ventriculitis management protocol: 11/26 (42.3%)<br>Mortality: 0/11 (0%) |
| Administration route | Not specified: 10/26 (38%)<br>Rickman reservoir: 14/26 (54%)<br>Omayya reservoir: 1/26 (4%)<br>Direct needle puncture: 1/26 (4%) |

**Table 3.**
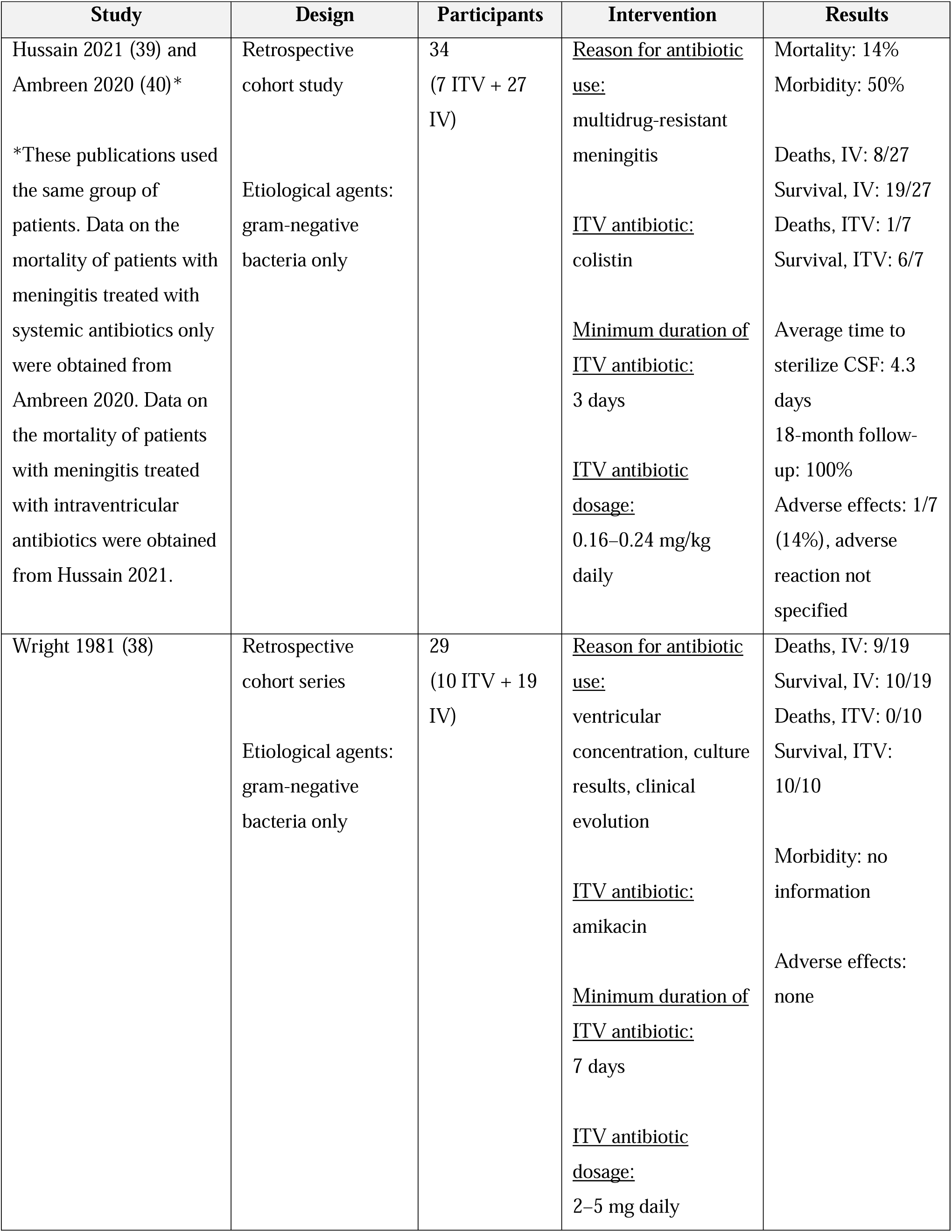
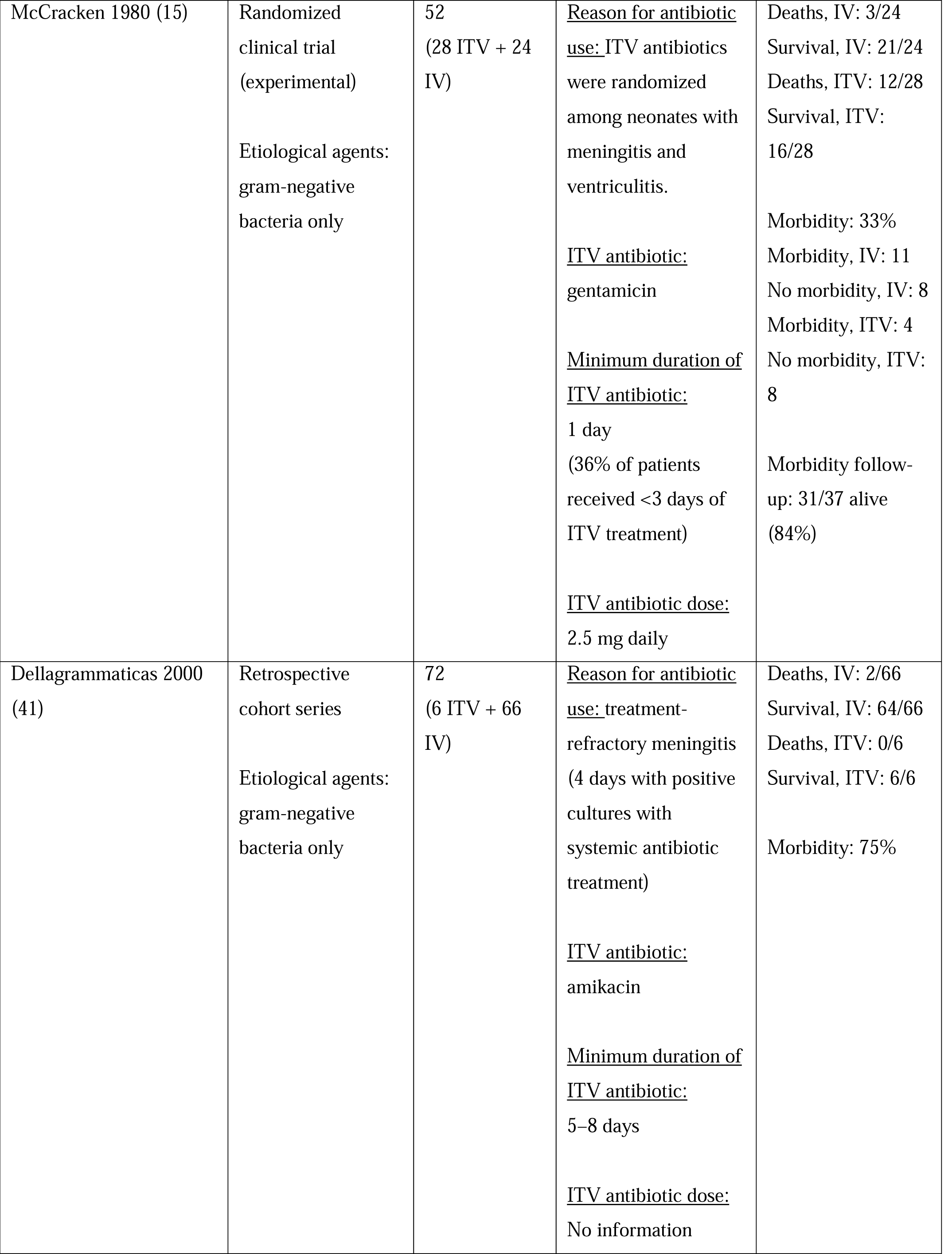
Description of the studies included in the meta-analysis of intraventricular antibiotics in pediatrics. ITV: intraventricular antibiotic; IV: systemic intravenous antibiotic.

**Table 4.**
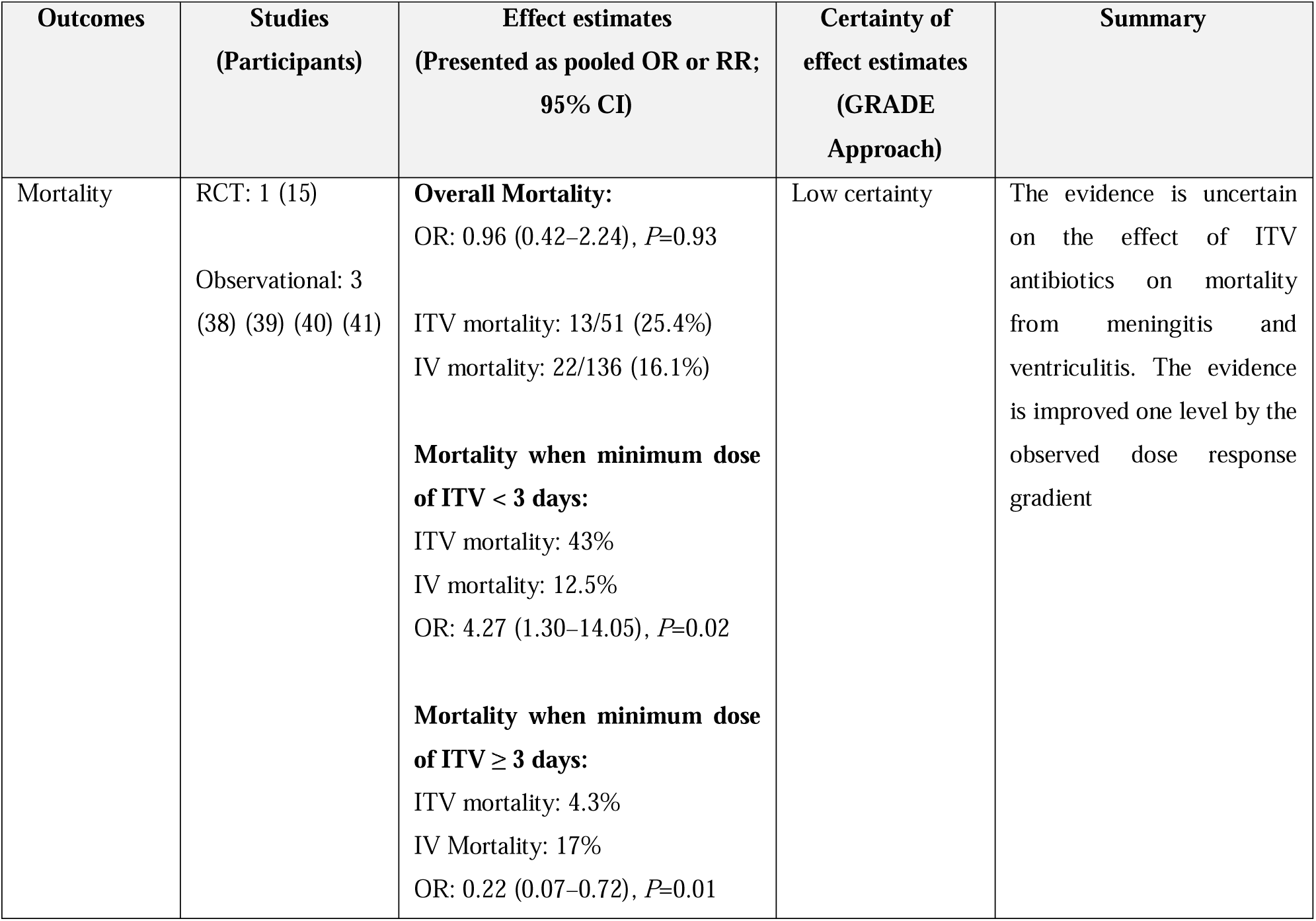
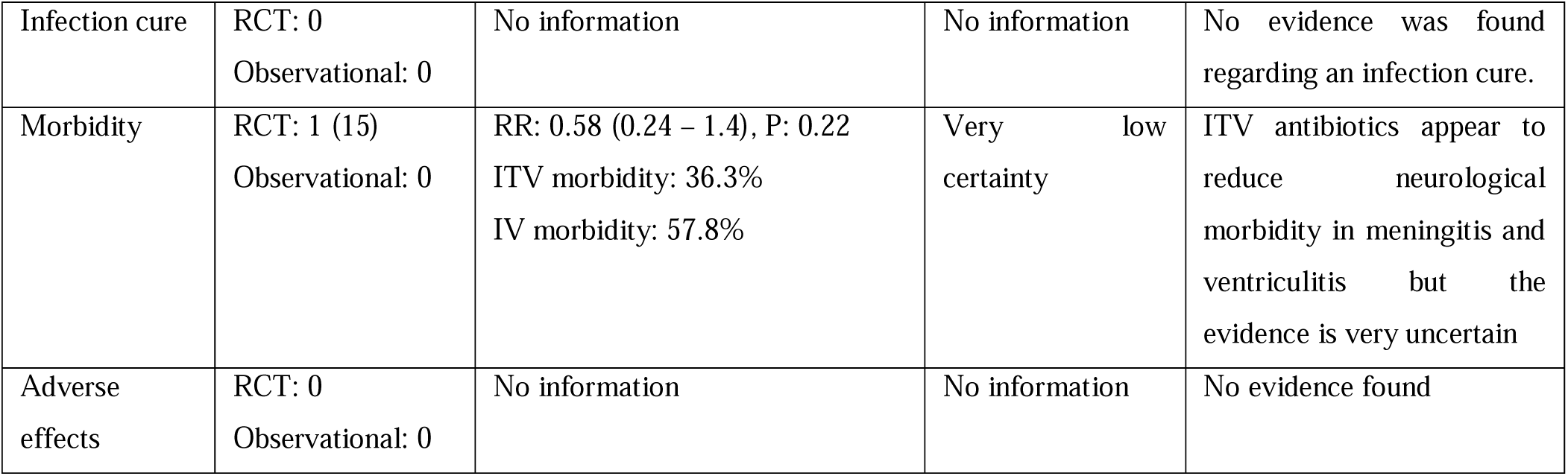
Summary of meta-analysis results. ITV: intraventricular antibiotics (plus systemic antibiotics); IV: systemic intravenous antibiotics only; RCT: Randomized clinical trial.

Regarding mortality, we included four studies and obtained an estimated effect in the meta-analysis, yielding an OR of 0.96 (0.42–2.24) and test for general effect of Z=0.09 (*P*=0.93) (Figure 4); therefore, no significant difference in mortality was observed with the use of intraventricular antibiotics in neonates. The certainty of this estimate was low (downgraded due to a high risk of bias, imprecision, and publication bias; the evidence was improved by one level due to the observed dose-response gradient, indicating a protective effect in studies where the minimum dose used was 3 days (Figures 2 and 3).

**Figure 2.**
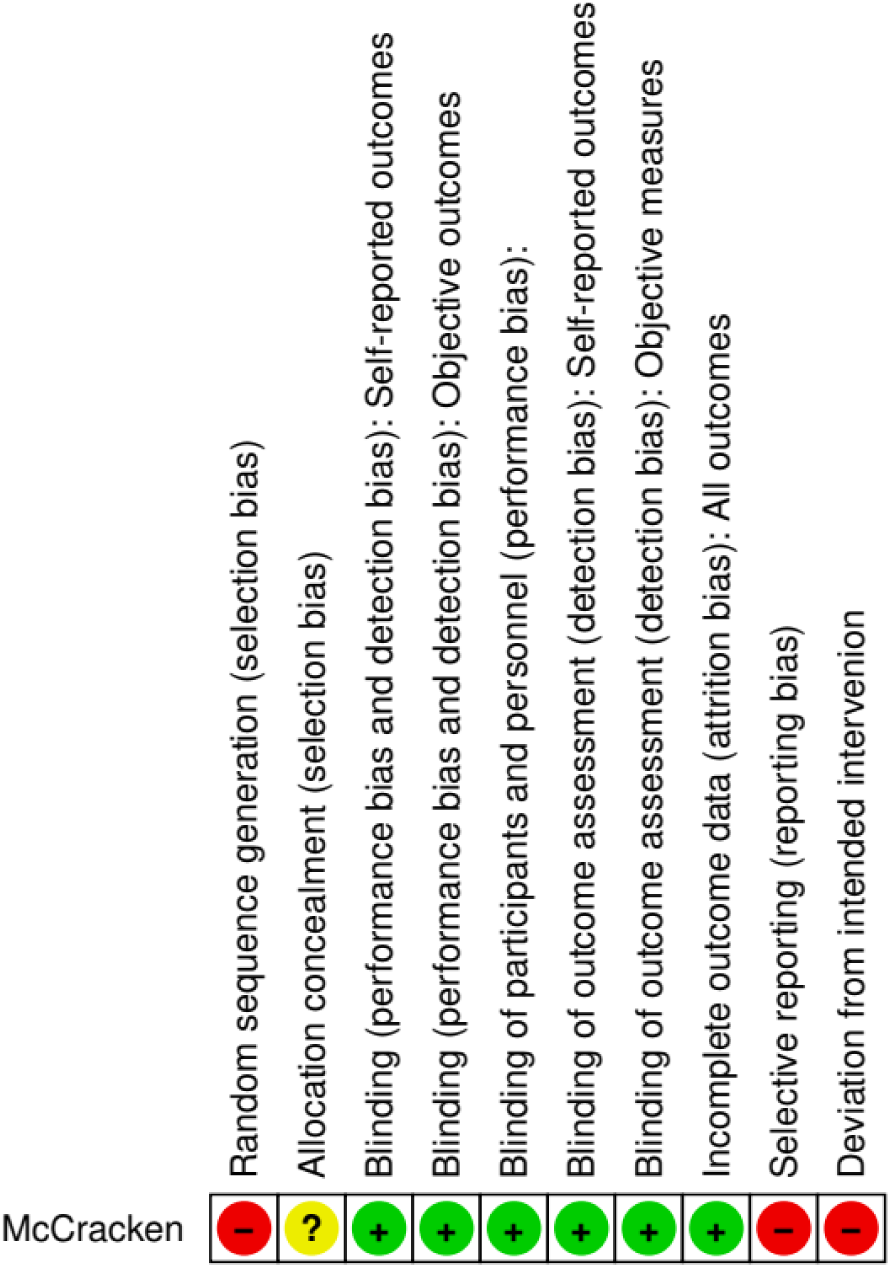
Risk of bias summary of randomized clinical trials of intraventricular antibiotics in pediatrics: review authors’ assessment of the risks of bias for each included study, using the ROBINS 2 tool. Graphic created with RevMan web.

**Figure 3.**
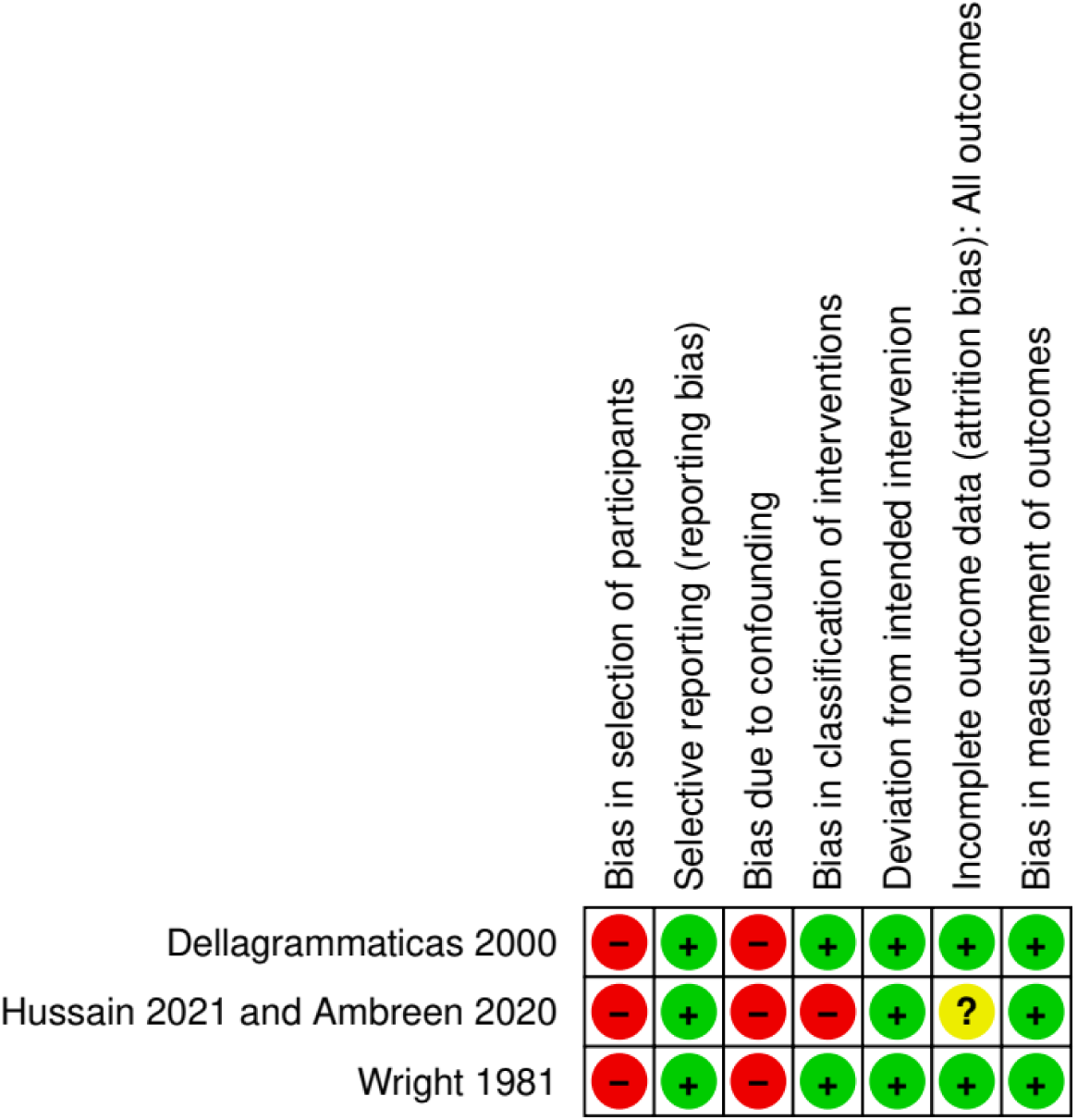
Risk summary of observational studies of intraventricular antibiotics in pediatrics: review authors’ assessment of the risks of bias for each included study, using the ROBINS 1 tool. Graphic created with RevMan web.

**Figure 4.**
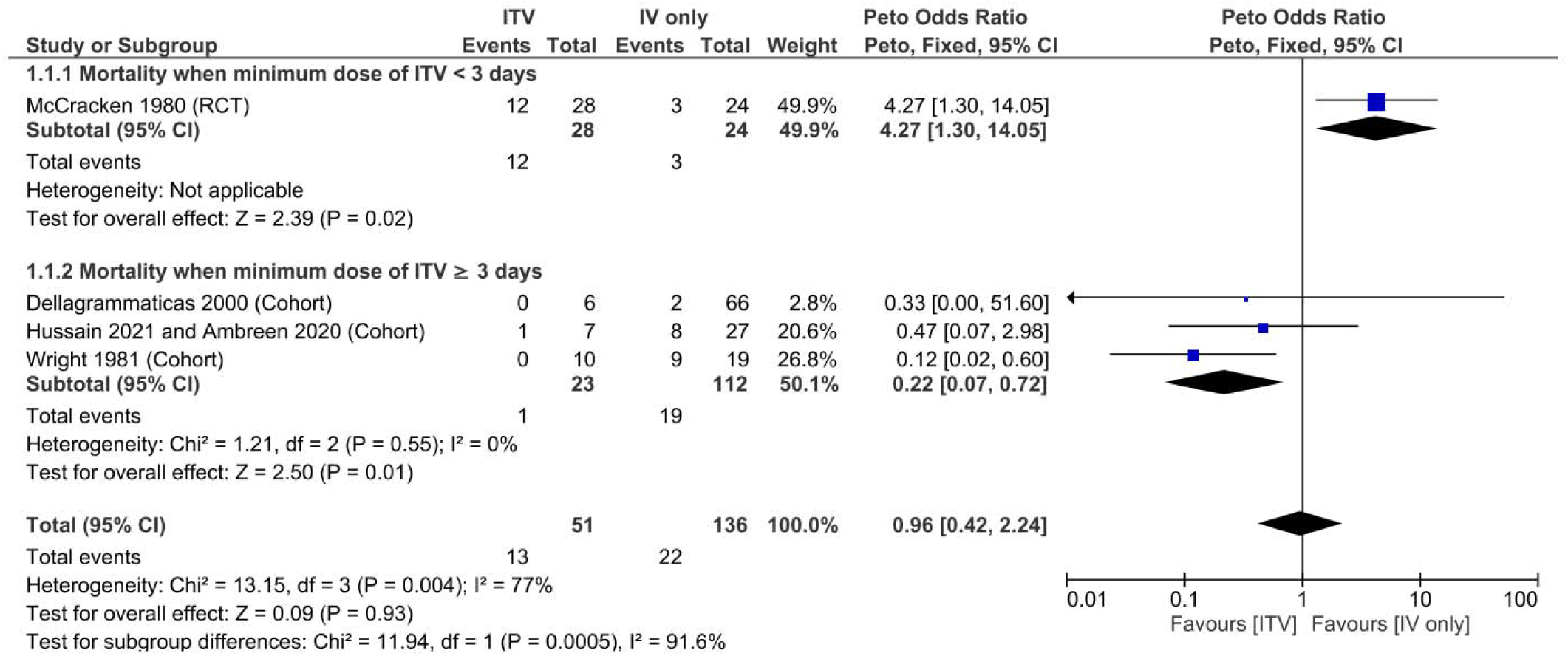
Meta-analysis of mortality of neonates treated with intraventricular antibiotics vs. systemic antibiotics only. ITV: intraventricular antibiotic (plus systemic antibiotic), IV: systemic intravenous antibiotics only.

The overall mortality estimate showed a high heterogeneity (*I*^2^=91.6). The heterogeneity could be explained by performing a subgroup analysis according to the minimum dose of intraventricular antibiotic used in the studies (studies with doses ≥ 3 doses of intraventricular antibiotic without heterogeneity: *I*^2^=0%) or by the type of study (experimental and non-experimental). Given the low quality of all included studies, our analysis suggests that the latter option is less likely (“Risk of bias assessment”).

In the subgroup of studies where the minimum duration of intraventricular antibiotics was ≥ 3 days, pooled mortality was significantly lower [4.3% vs 17%, OR=0.22 (0.07–0.72, *P*=0.01)].

Only one study was included for the analysis of neurological morbidity, with an estimated RR of 0.58 (0.24–1.4). The certainty of this estimate was very low (degraded due to the high risk of bias, imprecision, and publication bias) (Figure 2).

### Risk of bias assessment

#### Experimental study

Regarding the only randomized clinical trial (RCT) evaluated^15^, the overall risk of bias was high and was analyzed using the Cochrane ROBINS 2 tool (RoB 2).^43^ The methodological aspects in which we found serious weaknesses were randomization, deviation from the planned intervention, and selective reporting (Figure 2).

Randomization of the RCT^15^ could have been severely compromised because significant differences in clinical characteristics between the study groups were observed (Table 5).

**Table 5.** Difference in the distribution of clinical characteristics between the study groups. These are differences in the absolute and relative number between each group obtained from the first table of the randomized clinical trial.^15^.

| Clinical features | Group A<br>(systemic antibiotics<br>only)<br>(n) | Group B<br>(systemic + intraventricular<br>gentamicin)<br>(n) | Distribution difference<br>between treatment groups |
| --- | --- | --- | --- |
| Female | 14 | 10 | 28.5% |
| Male | 10 | 18 | 44% % |
| Birth weight > 2500 g | 6 | 14 | 57% |
| Onset to therapy<br>(hours)<br>MEAN ± 2 SEM | 26 ± 12 | 41 ± 24 | 15 hours |

However, the risk of bias that could have most significantly influenced the results of this study is the deviation from the planned intervention. This is because, in the control group (receiving only systemic antibiotics), 10 of the 24 patients had their treatment changed (with the addition of intraventricular antibiotics) due to a lack of improvement in the characteristics of the cerebrospinal fluid, as confirmed by the same author in another publication months after the experimental clinical trial was published (in a response from the author of the study, to a letter to the editor sent by several researchers concerned about inconsistencies in this study).^13^

Deviation from the planned intervention in the RCT ^15^ could be the cause of the inconsistencies observed, since it reported a much higher mortality with the use of intraventricular antibiotics (Tables 4 and 6), but used suboptimal doses (< 3 days of intraventricular treatment) in 36% of patients (10 of 28) in the intraventricular antibiotic group. The RCT ^15^ stipulated the administration of a minimum dose of intraventricular antibiotic for three days, as it had previously been shown to be more effective.^35^

Table 6 presents the results of our analysis of the experimental study ^15^ based on the administered doses. According to the study:

**Table 6:**
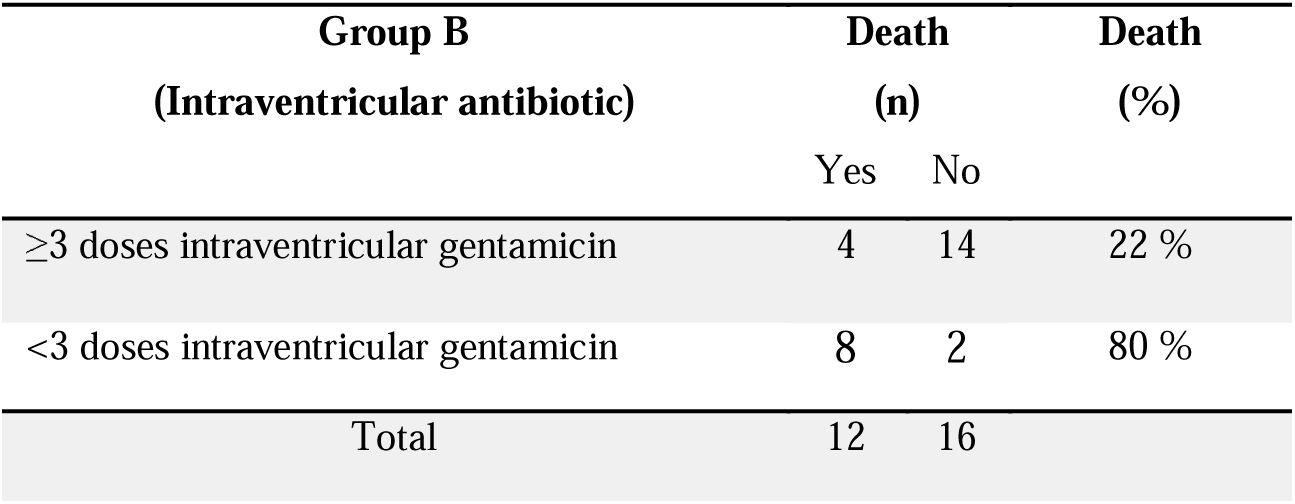
Results of the mortality stratified and dichotomized by antibiotic doses. Relative Risk = 0.27 (0.11–0.69), P = 0.003; absolute risk difference = 57.7%; relative risk difference = 72.5 %.

*“Of the 12 infants in group B (intraventricular antibiotic plus systemic antibiotics) who died, 6 received only 1 intraventricular dose of gentamicin, 2 received two doses, and 4 received 3 or more doses (mean 2.3 doses). In contrast, of the 16 survivors assigned to group B, only 2 received fewer than 3 doses and the average number of intraventricular doses administered was 4.7.”*

Thus, we rearranged (dichotomized) the results according to the minimum planned dose of intraventricular antibiotics ( 3 doses daily). ^15^ The results of our analysis are shown in Table 6. The difference in mortality between the minimum dose ( 3 doses of intraventricular antibiotics) was apparent.

Another risk of bias that we found in the reporting of the results in the RCT was due to the lack of analysis of the morbidity results in the original report. When we analyzed it in detail, we verified that with intraventricular antibiotics, a lower morbidity was achieved (36.3% vs. 57.8%) than with systemic antibiotics alone (Table 4).

#### Non-experimental studies

In the three observational studies included in the meta-analysis, ^38–41^ the overall risk of bias was high, as analyzed using the Cochrane ROBINS 1 tool (RoB 1).^44^ The methodological aspects in which we found serious weaknesses were confounding factors, classification of interventions, and participant selection (Figure 2).

Due to the non-randomized nature of the three observational studies and the lack of adequate evaluation of confounding factors, the risk of imbalance was high between the intervention groups.

Although the most recent observational study was published in two articles, ^39–40^ the data were generally consistent between publications and complemented each other. However, there is one piece of information that presents a high risk of information bias in intervention classification: the information on the mortality of the control group, which was not very clear and had to be deduced conservatively among the published data. We have written to the study authors to confirm this information and have not received any responses.

As the three studies were not randomized, and adjustment strategies were not designed to limit selection bias, we concluded that the three studies present a high risk of participant selection bias.

## DISCUSSION

In general, the quality of evidence for the use of intraventricular antibiotics in neonates in neurosurgery and pediatric studies is low, resulting in low certainty (Level C evidence).

### Research in Neurosurgery

Neurosurgical studies that are entirely observational have shown that the use of intraventricular antibiotics in ventriculitis generally results in a mortality rate of 5%. This figure is well below the average mortality rate of neonatal ventriculitis without the use of intraventricular antibiotics (37.3%). ^4,15,45^ However, this descriptive analysis should be interpreted as exploratory and subject to a high risk of bias.

No adverse effects have been reported in the neurosurgical literature regarding the use of intraventricular antibiotics in neonates. Significant morbidity occurred in 25% of the patients, which is a reduction compared to the 50% morbidity reported in cases of neonatal meningitis. ^4,6^ This could lead us to think that intraventricular antibiotics is a safe treatment; however, the important variables (adverse effects and morbidity) were rarely reported (only 25% reported morbidity and 32.2% reported the variable of adverse effects), which indicates a high risk of information and publication bias.

Most cases in which intraventricular antibiotics were used in neurosurgery studies were in patients with multidrug-resistant infections, those refractory to systemic antibiotic treatment, and almost exclusively in patients with ventriculitis associated with medical devices to treat hydrocephalus. In these cases, the risk of developing meningitis and ventriculitis is high, ranging between 6% and 30%. ^46,47^ The use of these devices in neurosurgery also increases the risk of multidrug-resistant infections, increasing the risk of death by up to 58.8%. ^48^ This scenario is common in neurosurgery, and intraventricular antibiotics have been a very useful therapeutic tool. In this review, we found that in neurosurgery patients, the average mortality when using intraventricular antibiotics with multidrug-resistant ventriculitis in our study was 10% and in refractory infections was 16%.

We found only one previous review that studied the use of intraventricular antibiotics in children in neurosurgery research; however, it only included those treated for multidrug-resistant infections. ^49^ This review included 10 neonates in addition to children over 1 month of age who used intraventricular antibiotics with a mortality rate of 20%, which is similar to the 10% mortality rate that we found in 30 neonates with multidrug-resistant ventriculitis.

Recently, three studies were published in which intraventricular antibiotics were used for ventriculitis prophylaxis in hydrocephalus surgeries in neonates. They managed to significantly reduce the incidence of infections, and no adverse effects or deaths were reported. ^50–52^

### Research in Pediatrics

In pediatric research on intraventricular antibiotics, although we used the term meningitis to refer to neuroinfections requiring the use of intraventricular antibiotics, ventriculitis actually coexists in most cases.^15,39^

The pediatric studies in general showed no difference in mortality in neonates with the use of intraventricular antibiotics compared to systemic antibiotics alone, with an OR of 0.96 (0.42–2.24, *P*=0.93). However, this result presents great heterogeneity (*I*^2^=91.6%), in that if we only considered the type of studies included in the meta-analysis, we could erroneously conclude that the higher mortality shown by the only experimental study is actually the true effect of the treatment.

However, as mentioned previously, this single experimental study presents a high risk of bias since there was a deviation from the planned treatment, as evidenced by the fact that 36% of the participants in the intraventricular antibiotic group received a lower dose (less than three doses) than the minimum established before starting the study, and an adequate analysis was not carried out (for example, analysis by intention to treat). This nullified the benefit that the randomization of the participants and treatments should have provided. In fact, in this study ^15^ we found that the greatest protective factor against death was presented by a duration of intraventricular antibiotics ≥ 3 days, with Relative Risk = 0.27 (0.11–0.69, P = 0.003), (Table 6).

The loss of randomization in the experimental study ^15^ is evident in the significant imbalance in clinical characteristics between both study groups (Table 5), and is most evident because of a marked difference in the time of initiation of intraventricular antibiotic treatment (average difference of at least 15 h compared to the start of treatment) in the control group with respect to symptom onset.

The results of the 1980 experimental study ^15^ were unexpected, especially for pediatricians who had already successfully used intraventricular antibiotics in neonates to treat meningitis and ventriculitis. ^10–13,38,53^ Furthermore, some experts have suggested that the higher mortality from intraventricular antibiotics in this experimental study could have been due to insufficient doses. ^13,53^ Recently, the guidelines of The Infectious Diseases Society of America on nosocomial meningitis and ventriculitis also questioned the results of the experimental study on the mortality of intraventricular antibiotics ^15^ due to insufficient doses, and the guidelines consider that intraventricular antibiotics are an option in neuroinfections that responds poorly to systemic treatment. ^18^ (Figure 5)

**Figure 5.**
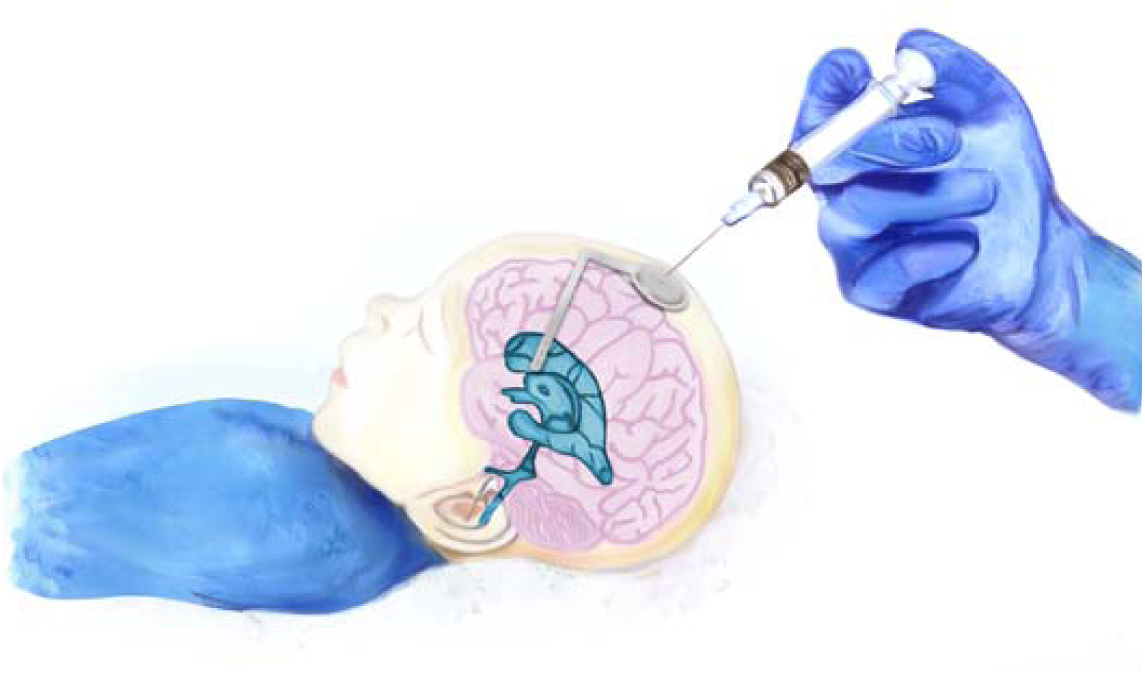
Diagrammatic representation of intraventricular antibiotic administration through a ventricular reservoir (used in pediatric studies).

The worrisome results of this experimental study ^15^ practically stopped the clinical use and research of intraventricular antibiotics in neonates in pediatrics, but not in neurosurgery, where they have continued to be used and expanded.

To elucidate the higher mortality associated with intraventricular antibiotics in neonates compared to conventional systemic treatment, the lead author of the experimental study ^15^ and other researchers reviewed cerebrospinal fluid samples from 21 patients in that same study to examine the concentrations of endotoxins, such as interleukin 1b and tumor necrosis factor. The objective was to find differences between patients treated with intraventricular antibiotics and those treated with systemic antibiotics alone. ^54^ A significant increase in the cytokine concentrations was observed in a group of samples from patients treated with intraventricular antibiotics. Although these results confirmed for the majority of the medical and scientific community the danger of using intraventricular antibiotics in neonates, it should be noted that they only included 21 patients in the study (of the 52 patients in the original experimental study), representing only 40% of the original sample (loss to follow-up of 60%); therefore, these results are not reliable due to the high risk of information bias. To be considered reliable, it is advisable to have studied at least 80% (loss to follow-up of less than 20%) of the original sample. ^55^

Furthermore, in the same study, cytokine concentrations were not correlated with the relevant clinical variables (mortality and morbidity); in particular, they were unable to find differences in mortality between patients treated with intraventricular antibiotics and systemic antibiotics alone (mortality: 20% vs. 18%). However, a lower morbidity was found among patients treated with intraventricular antibiotics (33% vs. 75%, *P*=0.19). ^54^

When analyzing the total administered doses of intraventricular antibiotics, we found a decreased mortality in studies in which a minimum dose of ≥ 3 days (mortality: 4.3%) of intraventricular antibiotic was administered as compared to those who did not use intraventricular antibiotics (mortality: 17%, *P*=0.01) (Table 4). This is an important decrease in mortality in neonatal meningitis compared with mortality in other studies, where the average mortality of neonatal meningitis was 20%. ^4,5,56–58^

Another explanation for the difference in mortality between the experimental and observational studies could be the varying indications for the use of intraventricular antibiotics in neonates. Observational studies administered intraventricular antibiotics based on the clinical condition of the patient, such as multidrug-resistant or treatment-refractory meningitis and ventriculitis, while in the experimental study, it was done randomly.

In the descriptive (non-comparative observational) pediatric studies (Table 2), we found a low mortality rate (7%), despite the fact that more than half (57.7%) of the patients were treated for meningitis refractory to conventional systemic treatment, which is usually caused by multi-drug-resistant bacteria. A previous study found that the mortality rate in multidrug-resistant neonatal meningitis was 58.8%, compared to 9.5% in infections caused by non-multidrug-resistant bacteria. ^48^ This comparison is not conclusive because these studies did not include a control group, but they can serve as exploratory studies, and more research could be conducted on this therapy.

A previous systematic review on intraventricular antibiotics in neonates was published in 2012; however, it only included a randomized experimental study. ^59^

## Conclusions

Considering the low quality of studies in pediatrics and neurosurgery, we conclude with a low level of certainty that intraventricular antibiotics may not have an effect on mortality in neonatal meningitis and ventriculitis. However, reduced mortality has been observed when a minimum duration of 3 days of intraventricular antibiotic treatment was used, particularly for multidrug-resistant or refractory infections. Better quality studies are needed to improve the quality of evidence and certainty regarding the use of intraventricular antibiotics for neonatal meningitis and ventriculitis.

## Data Availability

All data produced in the present study are available upon reasonable request to the authors

## Acknowledgment

We thank the technical support of the National Autonomous University of Nicaragua and particularly the professors from the Department of Statistics: Juan Ricardo Orozco (MSc) and José David García (MSc) for the statistical review. We also thank the independent review done by Drs. Dennis McDonnell (MD) and Abul Ariza Manzano (MD) for their invaluable contributions. Additionally, we appreciate the artistic work of drawing and diagrams made by Gloria Sarmiento Rodriguez (graphic designer). Lastly, we thank our families for their economic support, especially Marlene Valdivia, Alejandrino Perera, Elias Torres, and Tania Perera.

