## Supplement 1 for "Systematic review and meta-analysis of intraventricular antibiotics for neonatal meningitis and ventriculitis"

**SUPPLEMET 1**

**NEUROSURGERY CASE REPORTS**

| Study | n | Etiology | Antibiotic, dosage and duration | Infectious cure | Death | Note | Reason for use |
| --- | --- | --- | --- | --- | --- | --- | --- |
| Helgason 2022 | 1 | Methicillin-resistant Staphylococcus Aureus | Vancomycin | Yes  (1 of 1) = 100 % | No  (0 of 1)= 0% | -Adverse effects= 0%,  -No morbidity reported | Refractory infection |
| Bhat 2020 | 3 | Acinetobacter boumanii, Candida albicans, non Candida albicans | Colistin | Yes  (1 of 3)= 33 % | 1 of 3 =  33 % | Morbidity= 1 of 3 (33.3%) | Refractory infection |
| Pratheep 2019 | 1 | Acinetobacter boumanii | Tigecycline (3mg/day) and colistin (5 mg/day) for 2 weeks | 1 de 1 = 100% | 0 de 1 = 0 % | - Morbidity: not reported  - Adverse effects: 0% | Refractory infection |
| Joshi 2020 | 1 | Elizabethkingia meningoseptica | Vancomycin 10 mg /day for 2 weeks | 1 of 1 = 100 % | 0 of 1 = 0% | Morbidity: none  - Adverse effects: not reported | Multidrug-resistant infection (adjuvant treatment) |
| Piparsania 2012 | 1 | Acinetobacter boumanii | Polymyxin B | 1 of 1 = 100% | 0 of 1 = 0% | No morbidity or adverse effects | Multidrug-resistant infection (adjuvant treatment) |
| Nava-Ocampo 2006 | 1 | Enterococcus faecalis | Vancomycin 10 mg/day | 1 of 1 = 100 % | 0 of 1 = 0% | No reported morbidity or toxicity | Refractory infection |
| Laborada 2015 | 1 | Staphylococcus aureus | Vancomycin | 1 of 1 = 100 % | 0 of 1 = 0% | Study shows the variations of cytokines with atb. ITVa | Refractory infection |
| Greene 1983 | 1 | Citrobacter diversus | Amikacin  2 mg/day | 0 de 1 = 0 % | 0 de 1 = 0 % | Treatment with intraventricular amikacin failed but was later cured when trimetropim sulfa was added. | Multiresistant infection (adjuvant treatment) |
| Wirt 1979 | 1 | Serratia marcenscens | Amikacin 0.1 mg/ml CSF/day for 24 days | 1 of 1 = 100% | 0 of 1 = 0% | Study of 4 patients but only one is a neonate, whose results were reported. | Multidrug-resistant infection (adjuvant treatment) |
| Helms 1977 | 1 | E. coli | Gentamicin 2 mg/day for 8 days. | 1 of 1 = 100% | 0 of 1 = 0% | -Morbidity: none  -Adverse effects: not reported | Refractory infection |
| Matsunaga 2015 | 13 | Different species of staphylococcus, corynebacterium striatum | Vancomycin: between 5-20 mg/day | 13 of 13= 100% | 0 of 13= 0% | - Morbidity: not reported  - Adverse effects: 0% | Not reported |
| Yasidi 2018 | 1 | Enterobacter cloacae | Colistin 5 mg/day for 8 days | 1 of 1 = 100 % | 0 of 1 = 0% | No morbidity or adverse reactions reported | Refractory infection |
| Alaoui 2011 | 1 | Acinetobacter baumanii | Colistin 20,000 UI/kg/day | 1 pf 1 = 100 % | 0 of 1 = 0% | -Moderate or severe morbidity = not reported  -Adverse reactions= 0% | Multidrug-resistant infection (adjuvant treatment) |
| James 1984 | 18 | Retrospective cohort study. Infections after hydrocephalus surgery.  Etiological agents:  Gram positive = 72.2%  Gram negative = 27.7% | All patients were administered systemic and intraventricular antibiotics (which are not specified). | 17 of 18= 94.4% | Mortality = 1 /18 = 5.5% | -Morbidity = not specified  -Adverse effects: not reported | Multidrug-resistant infection (adjuvant treatment) |
| Nieto del Rincón 2000 | 2 | E. coli, S. epidermidis  Note: Postmortem diagnosis was not included. | It is not specified what antibiotics they used. |  | IVT deaths = 0/2 (0%)  Alive IVT = 2/2 (100%)  Deaths IV= 1/2 (50%)  Alive IV = 1/2 (50%) | Morbidity: 33%  Toxicity: not reported | Refractory infection |
| Tekgunduz 2015 | 1 | A. baumanii | Colistin, 10 mg/day for 21 days | 1 of 1= 100% | 0 of 1= 0% | Morbidity = not specified  Adverse effects: not reported | Refractory infection |
| Hiremath 2018 | 1 | A. baumanii | Colistin, 10 mg (125,000) /day for 3 days | 1 of 1= 100% | 0 of 1= 0% | Morbidity = not specified  Adverse effects: not reported | Multidrug-resistant infection (adjuvant treatment) |
| Mangi 1977 | 6 | Proteus mirabilis, pseudomona aeruginosa, E. Coli, | Gentamicin, from 5 to 42 mg for 1-3 days | 4 of 6 = 66 % | 2 of 6 = 33.3 % | Morbidity: no information  Adverse effects: none | Multidrug-resistant infection (adjuvant treatment) |
| Mehar 2012 | 1 | A. baumanii | Polymyxin B, 40,000 UI units alternate  day for 7 doses was given | Yes  (1 of 1) = 100 % | No  (0 of 1)= 0% | Morbidity: no information  Adverse effects: no information | Refractory infection |
