## Supplement 2 for "Systematic review and meta-analysis of intraventricular antibiotics for neonatal meningitis and ventriculitis"

**PEDIATRICS CASE REPORTS**

| Studies | n | Etiology | Antibiotic, dosage and duration | Infectious cure | Death | Note | Reason for use |
| --- | --- | --- | --- | --- | --- | --- | --- |
| Kaplan 1990 | 6 | K. pneumoniae, K.oxytoca, E.coli, Enterobacter cloacae, C.diversus | Aminoglycosides, Colistin.  Doses and duration were not stated. | 5 of 6 = 83% | 1 of 6 = 16.6 % | -Morbidity = no information  -Toxicity = no information | At the discretion of the treating physician |
| Rios 1978 | 1 | Flavobacterium meningosepticum | Ryfamicin 5 mg/day for 24 days | 1 of 1 = 100 % | 0 of 1 = 0 % | -Morbidity = none  -Toxicity = none | Multidrug-resistant infection (adjuvant treatment) |
| Lee 1977 | 16 | Flavobacterium meningosepticum, Escherichia coli, S. faecalis, Klebsiella aerogenes, proteous mirabilis, | Gentamicin (2 mg), Chloramphenicol, Rifampin (2-5 mg), Penicillin (5,000 IU), Erythromycin Base (10 mg) | 15 of16= 94% | 1 of 16 = 6 % | -Morbidity: 4 of 15 (survivors) = 26.6%  - Adverse effects: Temporary jaundice in 31.2% (5 of 16) who received rifampicin ITV. | Multidrug-resistant infection (adjuvant treatment) |
| Kaul 1978 | 3 | E. coli | Gentamicina (1mg/day)  And Gentamicin 1mg/day for 3 weeks | 3 of 3 = 100 % | 0 of 3 = 0 % | -Morbidity: 33.3%  -Adverse effects: not reported  IVT deaths = 0/3 (0%)  Live IVT = 3/3 (100%)  Deaths IV= 0 /2 (0%)  Alive IV= 2/2 (100%) | Refractory infection |
